# Workplace exposures may mask wildfire smoke-related exposure inequities and mortality

**DOI:** 10.64898/2026.02.04.26345584

**Authors:** Abas Shkembi, Sara D. Adar, Richard L. Neitzel, Marissa L. Childs

## Abstract

Millions of outdoor workers cannot avoid wildfire smoke, likely leading to inequalities in exposure and health risk. We characterized work-related exposure to wildfire PM_2.5_ for 3,108 contiguous US counties during 2006-2019. Despite experiencing less ambient exposure to wildfire PM_2.5_, counties with higher portions of non-Hispanic Black and Hispanic Americans experienced higher work-related exposure. We also find suggestive evidence that the effect of ambient smoke fine particulate matter (PM_2.5_) concentrations on all-cause mortality may differ by workplace exposure. These findings suggest that workplace exposures should be considered in wildfire smoke adaptation measures.

## Main

While the majority of the population can follow guidance to stay indoors with closed windows/doors on wildfire smoke days,^1^ the 45 million adults who regularly work outdoors in the United States (US) cannot.^2^ Outdoor workers are particularly susceptible to wildfire smoke due to direct exposures over long durations and intense exposure from increased metabolic rate, lack of ventilation, and potential co-exposure with other hazards. Current policy approaches to reduce exposures rely on personal protective behaviors as wildfire smoke can be exempted under the Clean Air Act, and the fluctuating concentrations of wildfire smoke allow for short-term behavioral changes that can lead to reduced exposures in the general population.^3^ Research has acknowledged the potentially greater exposure and health risks for outdoor workers^4–6^ and three states have enacted regulations to protect workers,^7^ but there are no comprehensive, nationwide estimates of people exposed to wildfire smoke through outdoor occupational exposure.^8^ Our lack of understanding of workplace exposures may mask wildfire smoke-related exposure inequities and human health impacts.

Racial and ethnic minority communities experience higher air pollution concentrations from all-source particulate matter in the US,^9^ yet studies of ambient particulate matter from wildfire smoke have observed the opposite—that some of these communities have lower concentrations.^10,11^ While previous studies have assumed that all populations within the same area experience wildfire smoke equally, racial and ethnic minority individuals are overrepresented in outdoor work such as farming, construction, and landscaping.^12^ This may lead to inequalities from workplace exposure that are not captured by comparisons of ambient concentrations alone.

Workplace exposure to wildfire smoke may also affect the health impacts experienced by individuals. We hypothesize that previously observed small and mixed existing evidence of wildfire-health risks^13^ could be partially explained by diverging behavioral patterns—i.e., since people who do not work outdoors avoid wildfire smoke by remaining indoors,^3^ while people who work outdoors cannot. These disparities between outdoor concentrations and personal exposures have importance for estimating the population-level health impacts of wildfire smoke. Analyses that relate ambient concentrations to adverse health outcomes will measure a combination of the experiences of people who do and do not work outdoors, resulting in a pooled overall effect that is likely weighted towards a smaller risk due to the larger, non-outdoor worker population in the US and their protective behaviors.

In this study, we quantified workplace exposures to wildfire smoke and explored the potential implications of differential exposure on racial and ethnic exposure inequities and mortality risk. We build on our previous approach characterizing work-related heat exposure to inform our estimates of work-related wildfire smoke exposure.^14^ First, we merged previously published estimates of ambient wildfire fine particulate matter (PM_2.5_) levels^15^ with employment counts and occupational characteristics (working outdoors, irregular hours) that influence wildfire smoke exposure to characterize annual work-related exposure to wildfire PM_2.5_ between 2006-2019 for outdoor workers among all 3,108 counties in the contiguous US. We focused on years prior to the COVID-19 pandemic due to changes in work environment and processes (e.g., who stays home, who works outdoors, availability of respirators) (see **Methods**). We then defined work-related wildfire smoke exposure as the number of outdoor worker-days per year where smoke PM_2.5_ exceeded 9 *μ*g/m^3^, based on the suggested threshold of the US National Institute for Occupational Safety and Health (NIOSH) to trigger protective actions by employers from all-source PM_2.5_ on wildfire smoke days (**Table S1**).^16^ To understand the implications of this occupational exposure for racial and ethnic inequities, we then compared the racial and ethnic composition of each county with the person-days exposed to ambient and work-related wildfire smoke. Lastly, we replicated a previous analysis of ambient wildfire smoke impacts on all-cause mortality,^17^ and stratified by work-related exposure to understand differential mortality effects of ambient wildfire PM_2.5_ (**Methods**). Using a quasi-Poisson fixed effects regression model, our approach accounted for any time-invariant confounders between counties, time-varying confounders within states, and time-varying confounding due to temperature and precipitation. We assessed these relationships for the entire population, rather than the working age population, because the US workforce is increasingly aging,^18^ child labor is common particularly in agriculture,^19^ and work-related illness from wildfire smoke could indirectly increase mortality risk for younger or older dependent family members (e.g., due to wage or health insurance loss).^20^

We found that between 2006-2019, there were an estimated 836 million person-days of exposure to wildfire PM_2.5_ ≥9 *μ*g/m^3^ among outdoor workers in the US (**Figure 1a**). On average, this equated to 16.3 of every 10,000 US workers being exposed every day. Exposures varied substantially by day, with 233 days of the 14-year period having >1 million workers exposed. Most exposures occurred in 2007 (20% of exposures), 2011 (15%) and 2018 (12%), years where a large population of the US was exposed to wildfire smoke.^15^ Farming, Fishing, and Forestry (average 75.8 per 10,000 workers) and Construction and Extraction (average 74.9 per 10,000 workers) occupations had substantially higher exposure rates than the national average (**Figure 1b**).

**Figure 1.**
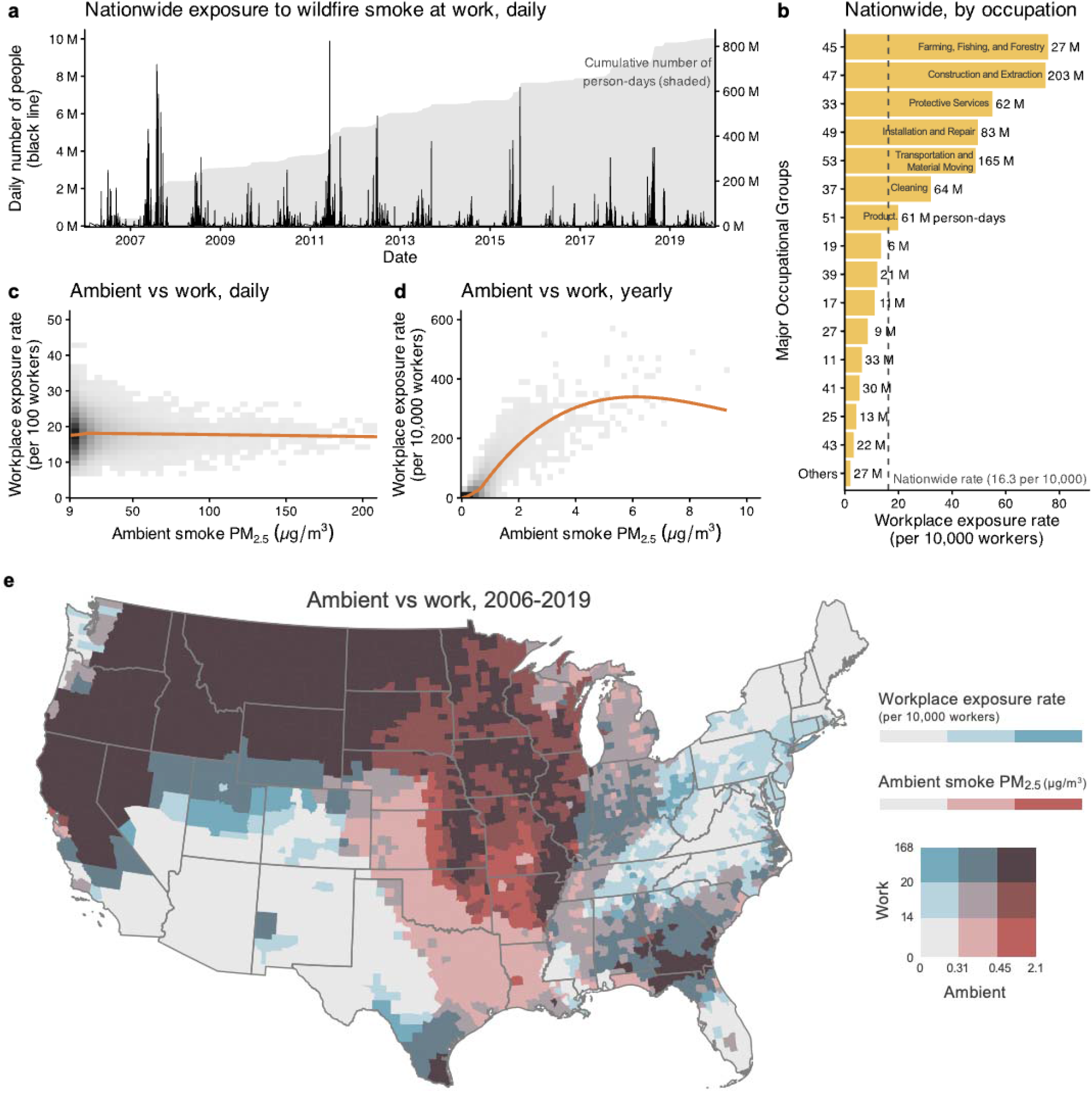
Exposure to wildfire PM_2.5_ smoke between 2010-2019 among outdoor workers in the contiguous US and comparison to ambient levels. **a** Daily number of people exposed (black line) and cumulative number of person-days exposed (grey shaded area). **b** Bar chart of the workplace exposure rate of wildfire smoke by major occupational group during 2006-2019 (bar labels present total number of person-days exposed). **c** The relationship between daily ambient smoke PM_2.5_ (*μ*g/m^3^) vs the daily rate of work-related exposure per 100 workers on county-days ≥9 *μ*g/m^3^ during 2006-2019. Shaded area reflects density of observations, with darker shades indicating more observations. Ambient smoke levels are truncated to ≤200 *μ*g/m^3^ to make lower values more visible since there are few days >200 *μ*g/m^3^. **d** The relationship between annual average, ambient smoke PM_2.5_ (*μ*g/m^3^) vs the annual rate of work-related exposure per 10,000 workers for every county-year during 2006-2019. Shaded area reflects density of observations, with darker shades indicating more observations. **e** Bivariate map of the daily rate of work-related exposure per 10,000 workers and average ambient smoke PM_2.5_ (*μ*g/m^3^) at the county-level during 2006-2019. Note: major standard occupational classification (SOC) group titles: 11 – Ma agement Occupations; 17 – Architecture and Engineering Occupations; 19 – Life, Physical, and Social Science Occupations; 25 – Educational Instruction and Library Occupations; 27 – Arts, Design, Entertainment, Sports, and Media Occupations; 33 – Protective Service Occupations; 37 – Building and Grounds Cleaning and Maintenance Occupations; 39 – Personal Care and Service Occupations; 41 – Sales and Related Occupations; 43 – Office and Administrative Support Occupations; 45 – Farming, Fishing, and Forestry Occupations; 47 – Construction and Extraction Occupations; 49 – Installation, Maintenance, and Repair Occupations; 51 – Production Occupations; 53 – Transportation and Material Moving Occupations.

Daily ambient wildfire PM_2.5_ smoke concentrations correlated poorly with total work-related exposures (**Figure 1c**). While annual average ambient wildfire PM_2.5_ smoke concentrations correlated more strongly with work-related exposure, increasing ambient levels at the tails (<0.5 or >4 *μ*g/m^3^) were not strongly correlated with higher work-related exposure (**Figure 1d**). When averaging across 2006-2019, **Figure 1e** illustrates there are many counties with the highest of ambient wildfire PM_2.5_ but the lowest workplace exposure rates. This suggests that characterizing workplace exposures can provide information about wildfire smoke exposure beyond the information ambient levels provide.

Ambient measures also masked racial and ethnic exposure inequities to wildfire PM_2.5_ smoke (**Figure 2a**). While counties with higher proportions of non-Hispanic White individuals in the population were more exposed to ambient wildfire PM_2.5_ on average, they were less exposed to wildfire PM_2.5_ at work. Conversely, while counties with higher proportions of non-Hispanic Black and Hispanic individuals were less exposed to ambient wildfire PM_2.5_, both were more exposed at work, particularly counties with higher portions of Hispanic individuals.

**Figure 2.**
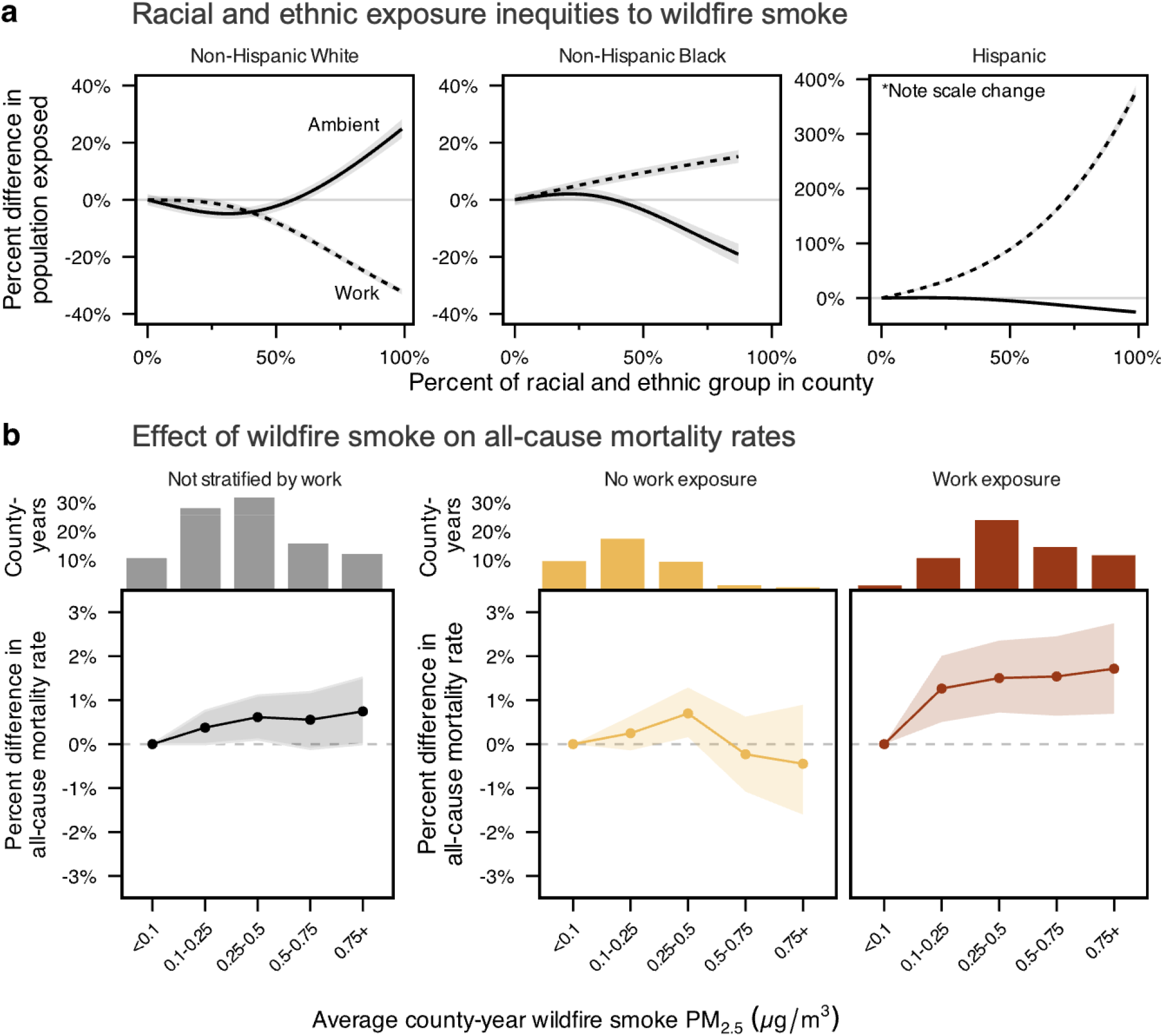
Workplace exposure to wildfire smoke PM_2.5_ may mask wildfire-related exposure inequities and all-cause mortality risk. **a** County-year relationship between racial and ethnic composition of a county and the rate of ambient (solid line) and workplace (dashed line) wildfire PM_2.5_ exposure during 2006-2019. Percent differences are calculated relative to counties with 0% of the population identifying as the respective racial and ethnic group. **b** County-year association from panel fixed effects quasi-Poisson regression models between average, ambient smoke PM_2.5_ and all-cause mortality rates between 2006-2019 among all counties (left panel), and among counties with no (0 exposure-days per 10,000 workers), or any (>0 exposure-days per 10,000 workers) workplace exposure (right two panels).

Differential workplace exposure can lead to detectable differences in all-cause mortality rates (**Figure 2b; Table S2**). Consistent with previous studies,^17,21^ we found an increasing trend between annual ambient wildfire smoke PM_2.5_ and all-cause mortality rates when all counties were pooled together, although the effects at higher averages >0.5 *μ*g/m^3^ were not statistically significant. Among county-years with no workplace exposure (**Figure S1**), mortality risk peaked at 0.25-0.5 *μ*g/m^3^ (0.7% higher mortality rate, 95% CI: 0.2 to 1.2%) relative to no/minimal ambient smoke PM_2.5_ (<0.1 *μ*g/m^3^), with no increase in mortality risk for ambient annual concentrations >0.5 *μ*g/m^3^. This effect is compatible with the negative effects of smoke on emergency department visits at high daily concentrations observed previously,^22^ suggesting those without workplace exposure could be taking measures to avoid wildfire smoke.^3,13^ In contrast, county-years with workplace exposure to wildfire smoke (**Figure S2**) displayed statistically significantly higher all-cause mortality rates (1.2 to 1.7%) across all smoke PM_2.5_ concentrations relative to no/little ambient PM_2.5_, consistent with our hypothesis that outdoor workers cannot avoid exposure.

Collectively, these findings suggest estimates of ambient wildfire smoke concentrations alone can mask wildfire smoke exposure inequities and all-cause mortality driven by work-related exposures. The observed inequities indicate that racial and ethnic minority communities, particularly Hispanic, may disproportionately bear the health burden of wildfire smoke, although further research is needed to investigate potential health disparities. The differential mortality burden suggests that wildfire smoke adaptation measures may be beneficial in areas with higher workplace exposure to wildfire smoke.

There are several factors to consider when interpreting our results. We may have mischaracterized work-related exposure in some counties, although we made specific analytic decisions that avoid strong bias in either direction (see **Appendix A**). This analysis does not consider other individual characteristics, like income or housing quality, which affect individual exposures. Future studies should consider incorporating individual-level information. While our regression approach replicates previous, plausibly causal estimations of wildfire smoke and mortality,^17^ there may be additional, unmeasured time-varying confounders that could bias our observed effects (e.g., short-term labor “shocks”). However, these confounders would need to be correlated with wildfire smoke PM_2.5_ within counties. Our observed differential mortality effect is sensitive to the threshold that defines work-related exposure (9 *μ*g/m^3^), however, this threshold was guided by available recommendations.^16^ This makes it difficult to disentangle the contribution of workplace exposure or the higher exposure levels more generally to the observed differential effect.

Regardless of these limitations, the differential mortality effect is plausible for several reasons. Working outdoors is a large risk factor for wildfire smoke exposure due to potential prolonged exposure (>8 hours/day). Outdoor workers also have limited options to avoid and control exposure to wildfire smoke. The few that exist, such as placing workers in semi-permanent structures, implementing air filtration systems, rotating work schedules, altering work intensity, and providing respirators,^16^ were unlikely to have been implemented in many workplaces as of 2019 due to a lack of workplace regulations. Only three states (California, Oregon, and Washington) have since implemented regulations despite the increased geographic and temporal spread of wildfire smoke in the 2020s.^23^ The lack of sufficiently protective regulations and the differential mortality impacts highlight the need to protect outdoor workers and consider occupational health in wildfire smoke adaptation measures in order to reduce the population health burden of wildfire smoke.

## Acknowledgements

The authors would like to acknowledge the work by Childs et al. (daily smoke data); Bureau of Labor Statistics (OEWS data); Department of Labor (O*NET data); the US CDC National Center for Health Statistics (mortality data); and Kyle Walker (tidycensus R package) for making data feely available for researchers to use. This study would not have been possible without their efforts. Research reported in this publication was not supported by any funding sources.

## Author Contributions

AS and MLC conceptualized the research; all authors designed the analysis; AS performed the research and analyzed data; AS drafted the first paper and all authors revised the paper.

## Data Availability

Yearly county breakdowns of workplace wildfire smoke exposure rates are available at *[note: will be available near publication]*. For daily exposure rate estimates, contact Abas Shkembi. All data underlying the analysis are publicly available. Major employment counts and sociodemographic compositions are available from the ‘tidycensus’ R package (https://cran.rproject.org/web/packages/tidycensus/index.html). Daily, ambient wildfire smoke levels were previously published by Childs et al. (https://github.com/echolab-stanford/daily-10km-smokePM). Metropolitan/nonmetropolitan-level detailed employment counts are available by the Bureau of Labor Statistics Occupational Employment and Wage Statistics (https://www.bls.gov/oes/tables.htm). Workplace characteristics are made publicly available from version 24.1 of the Occupational Information Network (https://www.onetcenter.org/db_releases.html). Mortality data are made publicly available by the CDC National Center for Health Statistics on the WONDER platform (https://wonder.cdc.gov/deaths-by-underlying-cause.html).

## Code Availability

Replication code required to reproduce the results are maintained in the following GitHub repository: *[note: will be available near publication]*

## Methods

Building on previous work characterizing work-related heat exposure,^14^ we characterized the number of person-days of wildfire PM_2.5_ exposure among outdoor workers in the contiguous US. We integrated previously published, county-level, daily estimates of wildfire smoke fine particulate matter (PM_2.5_) from 2006 to 2019^15^ with county-level estimates of employment counts by detailed occupational groups (760 civilian occupations, as defined by the 2010 US Bureau of Labor Statistics [BLS] Standard Occupational Classification [SOC] system, **Figure S3 and Table S3**).^24^ For each detailed SOC code, we considered two job characteristics that have data publicly available and that could influence exposure: (a) outdoor work and (b) irregular work hours, both derived from version 24.1 of the 2019 Occupational Information Network (**Figures S4-6**).^25^ For each day and county between 2006-2019, we estimated the number of all workers who work outdoors in a given detailed SOC group within each county that are potentially exposed to smoke PM_2.5_ levels >9 *μ*g/m^3^, a protective threshold suggested by the NIOSH.^16^ More details on how the measures used and the approach to derive these estimates are provided in **Appendix A**. These estimates were aggregated both spatially (e.g., nationally), occupationally (e.g., major SOC groups), and temporally (e.g., yearly and across 2006-2019) to be compared to ambient exposure levels, but our main indicator of interest for racial and ethnic inequalities and differential mortality effects was the annual, county-level rate of work-related wildfire PM_2.5_ exposure, expressed as the daily number of exposure events per 10,000 worker-days (**Figure S7**).

To investigate whether workplace exposures mask racial and ethnic wildfire smoke inequities, we ran two Poisson, generalized additive models to assess the relationship between the racial and ethnic composition in each county and year (the percent non-Hispanic White, non-Hispanic Black, or Hispanic) with the number of person-days exposed to (i) ambient or (ii) workplace wildfire smoke. We use person-days exposed as the outcome for ambient wildfire smoke, rather the more conventional average concentration level, to facilitate comparison between the workplace wildfire smoke metric which is measured in person-days. Both models accounted for temporal and spatial autocorrelation, and the workplace model additionally adjusted for the population unlikely to be working, and thus unable to have work-related exposure (see **Appendix B**).

To investigate whether wildfire smoke mortality risk differs by workplace exposure, we then fit a quasi-Poisson, panel fixed effects model using county-year data to assess the relationship of the joint exposures to ambient and work-related wildfire PM_2.5_ levels with all-cause mortality. This approach replicates the analytical design and model specifications of a previous wildfire smoke-mortality analysis using similar data (**Figure S8**).^17^ We obtained all-cause mortality counts for each county for all contiguous US counties between 2010-2019 from the National Center for Health Statistics.^26^ We included county and state-year fixed effects, natural cubic splines with 5 degrees of freedom for mean county-year temperature and precipitation,^27,28^ and an offset term of county-year total population. Additional details of the regression model, justification of modeling choices, and sensitivity analyses (**Figure S9**) are provided in **Appendix C**.

