## Supplemental Materials for "Workplace exposures may mask wildfire smoke-related exposure inequities and mortality"

**Table of Contents**

**List of Supplementary Tables**

- Supplemental Table S1-3

**List of Supplementary Figures**

- Supplemental Figures S1 - S9

**Supplemental Methods and Results**

- Appendix A. Characterizing workplace exposure to wildfire PM_2.5_
  - Determining exposure to wildfire smoke
  - Daily wildfire smoke exposure data
  - Detailed workplace characteristic data
  - Detailed employment count data by county
  - Exposure estimation
  - Poststratification using major employment count data by county
  - Assessment of analytical decisions on estimation bias
- Appendix B. Relationship between racial and ethnic composition and ambient and work-related exposure to wildfire PM_2.5_
  - Data sources
  - Model specifications
- Appendix C. Relationship between ambient wildfire PM_2.5_, workplace exposure, and mortality relationship
  - Data sources
  - Model specifications
  - Sensitivity analysis

**List of Supplementary Tables**

**Supplementary Table S1.** NIOSH exposure control categories (ECC) by Air Quality Index groupings as defined in Table 4-1 in the draft 2024 NIOSH Hazard Review “Wildland Fire Smoke Exposure Among Farmworkers and Other Outdoor Workers”.^1^

**Supplemental Table S2.** Percent change (95% confidence interval) between average, ambient smoke PM2.5 and all-cause mortality rates between 2006-2019 among all counties (pooled), among counties with no (0 exposure-days per 10,000 workers), or among counties with any (>0 exposure-days per 10,000 workers) workplace exposure from panel fixed effects quasi-Poisson regression models of county-years. Effects visualized in Figure 2b.

**Supplemental Table S3.** List of each variable used to construct work-related wildfire smoke exposure rates across three steps (Step 1: Data integration; Step 2: Exposure estimation; and Step 3: Poststratification).

**List of Supplementary Figures**

**Supplemental Figure S1.** Annual average ambient wildfire smoke PM_2.5_ levels (µg/m^3^) among county-years with no work-related exposure to wildfire smoke. States with white border have occupational regulations on wildfire smoke as of 2025.

**Supplemental Figure S2.** Annual average ambient wildfire smoke PM_2.5_ levels (µg/m^3^) among county-years with any work-related exposure to wildfire smoke. States with white border have occupational regulations on wildfire smoke as of 2025.

**Supplemental Figure S3.** Flowchart illustrating data inputs, statistical methodology, and output of the workplace exposure rate per 1,000 workers. BLS OEWS: Bureau of Labor Statistics Occupational Employment and Wage Statistics; O*NET: Occupational Information Network; US ACS: United States American Community Survey.

**Supplemental Figure S4.** Histogram by detailed SOC group (n=760) of the proportion of work-year outdoors

**Supplemental Figure S5.** Yearly, likely number of outdoor workers by county. States with white border have occupational regulations on wildfire smoke as of 2025.

**Supplemental Figure S6.** Bar chart by detailed SOC group (n=760) of irregular working hours and could work on the weekends (value = 1)

**Supplemental Figure S7.** Yearly, average rate of work-related wildfire PM_2.5_ exposure ≥9 µg/m^3^ per 10,000 workers by county. States with white border have occupational regulations on wildfire smoke as of 2025.

**Supplemental Figure S8.** Sensitivity analysis comparing original effect estimates (top to bottom) reported by Qiu et al. (using restricted-use mortality data without suppressed mortality counts), replicating Qiu et al.’s model (using publicly available mortality data with suppressed mortality counts and different data sources for model covariates), and truncating the replicated model to >0.75 µg/m^3^.

**Supplemental Figure S9.** Sensitivity analysis comparing effect estimates from differences in handling suppressed mortality values among county-years with <10 deaths. Top to bottom: imputing suppressed values to 4.5 (mid point of 0-9 deaths; main model); imputing suppressed values to 0; imputing suppressed values to 9; dropping county-years with suppressed values.

**Supplemental Methods and Results**

**Appendix A. Characterizing workplace exposure to wildfire PM_2.5_**

Building on a previous characterization of work-related heat exposure in the US,^2^ we characterized work-related wildfire smoke exposure for every county-year from 2006 to 2019 in the continental US. A simplified illustration of data inputs, statistical methodology, and data output required to characterize workplace is presented in **Supplemental Figure S3.** A detailed list of each variable used is presented in **Supplemental Table S3**.

**Determining exposure to wildfire smoke.** No nationwide regulations for the protection of outdoor workers during wildfire smoke PM_2.5_ exposure events exist in the US. The National Institute of Occupational Safety and Health (NIOSH), a governmental research agency that is part of the United States Centers for Disease Control and Prevention that sets recommendations for the regulatory Occupational Safety and Health Administration, developed draft guidelines for assessing workplace exposure to wildfire smoke in 2024. These guidelines were not finalized as of October 2025. We used guidelines set in Table 4-1 to determine exposure to wildfire smoke.^1^

The draft NIOSH guidelines group workers into six Exposure Control Categories (ECCs) based on Air Quality Index (AQI) groupings during wildfire events.^1^ The ECC groups are summarized in **Supplemental Table S1**. These ECCs are based on all-source PM_2.5_ rather than wildfire-specific PM_2.5_, however, our approach here utilizes wildfire-specific PM_2.5_ levels. Our rationale for our approach is that we wanted to assess the population exposed specifically to wildfire smoke, which NIOSH’s approach cannot tease out. We only considered exposure among wildfire PM_2.5_ levels exceeding 9 µg/m^3^, for two reasons. First, this is the threshold that NIOSH recommends employers to take control action to immediately protect workers from wildfire smoke.^1^ Second, acceptable exposure levels in workplaces are typically much higher than in communities, so assessing exposure <9 µg/m^3^ may not reflect levels high enough to prompt employers to provide workplace controls or Personal Protective Equipment (PPE) such as respirators.

Specific occupations, such as wildland firefighters, are the most burdened occupation during wildfire events; however, the aim of our study was to characterize downwind smoke exposure for all outdoor workers, not just primary exposure due to fighting wildfires. Overall, we analyzed exposure from 2006 up until 2019 because in 2020, the COVID-19 pandemic altered which workers continued to go to work and whether they are required to work outdoors. Changes to outdoor work beginning in 2020 could not be adequately captured with the workplace information data used in this study, so we limit our estimated exposure to 2019.

**Daily wildfire smoke levels.** We extracted daily, ambient wildfire PM_2.5_ estimates previously developed by Childs et al. between 2006-2019 for every county in the contiguous US.^3^ No data processing/cleaning was performed on these data for the purposes of this study. Briefly, these estimates were developed by Childs et al. by identifying days when smoke was overhead based on satellite imagery to calculate anomalies in station-based ground PM_2.5_ above the median on non-smoke days to define smoke PM_2.5_. Using information on meteorological factors, fire variables, aerosol measurements, land use, and elevation data, Childs et al. trained a model on the station-based smoke PM_2.5_ to be extrapolated to 10km x 10km grids of daily smoke PM_2.5_ across the contiguous US. Daily county estimates are population-weighted aggregates of these 10km x 10km gridded estimates.

**Detailed workplace characteristic data.** To determine whether a given occupation may be exposed to wildfire smoke, we leveraged data by the detailed, 2010, US Standard Occupational Classification (SOC) codes in the 2019 Occupational Information Network (O*NET, version 24.1).^4^ The O*NET database is made publicly available and developed by the US Department of Labor. The O*NET database provides estimates of work characteristics using self-reported data collected by the Department of Labor. The data are provided by detailed 2010, O*NET-SOC codes. The 9-digit, detailed O*NET-SOC codes were converted to 7-digit, detailed BLS SOC codes by aggregating the first 7 digits of the O*NET-SOC codes and taking the average value by detailed SOC code. Specifically, we determined the probability a detailed SOC group may be working outdoors and whether a detailed SOC group typically works on the weekend.

*Probability of outdoor work.* We used the continuous “Frequency Required to Work Outdoors, Exposed to Weather” work context variable (element ID #4.C.2.a.1.c). Specifically, workers responded to the following question: “How often does this job require working outdoors, exposed to all weather conditions?”. Possible responses ranged from “Never” to “Everyday”. This estimate is provided on a scale of 0-100, which was rescaled to 0-250 to reflect the standard, 250-day work-year, as previously done by our study team.^5^ We then converted the estimate back onto a scale of 0-1 to reflect the proportion of the standard work-year that a worker is working outdoors (see **Supplemental Figure S4 and S5**).

*Irregular work hours.* We used two variables to determine whether a detailed SOC group might work beyond a standard “9-5”, weekday job, as additional work on the weekend could increase exposure incidence for some jobs. First, we used the continuous “Duration of Typical Work Week” work context variable (element ID #4.C.3.d.8). Specifically, workers responded to the following question: “Number of hours typically worked in one week”. Values ranged from 0-100. Possible responses were from “Less than 40 hours” (value of 0), “40 hours” (value of 50), and “More than 40 hours” (value of 100). We assigned a binary value of whether a detailed SOC group works more than 40 hours if the value was >50. Next, we used the continuous “Work Schedules” work context variable (element ID #4.C.3.d.4). Specifically, workers responded to the following question: “How regular are the work schedules for this job?”. Values ranged from 0-100. Possible responses were from “Regular (established routine, set schedule)” (value of 0), “Irregular (changes with weather conditions, production demands, or contract duration)” (value of 50), and “Seasonal (only during certain times of the year)” (value of 100). We assigned a binary value of whether a detailed SOC group works irregularly if the value was >25. If either the first value was >50 or the second value was >25, we considered the detailed SOC group as “weekend workers” (see **Supplemental Figure S6**).

**Detailed employment count data by county.** We extracted 2019 metropolitan/nonmetropolitan area employment counts of detailed Standard Occupational Classification (SOC) codes from the US Bureau of Labor Statistics (BLS) Occupational Wage and Employment Statistics (OEWS).^6^ Metropolitan areas are counties grouped together based on common economic activities and commuting patterns around a city. Thus, some metropolitan areas are a single county, while others are a combination of a few counties. Nonmetropolitan areas are larger groups of many counties that do not have economic activity around a major city. We assumed that the spread of detailed SOC codes was the same for the counties within a metro/nonmetro area. These detailed SOC employment counts are not developed to be assessed in a time-varying manner, as the OEWS aggregates data across many years to develop their estimates. Thus, we used 2019 data to span across the years of analysis in this study (2006-2019). In 2018, SOC codes were updated from the 2010 SOC code version. We crosswalked all data to 2010 SOC codes since the majority of data used in this study used the 2010 SOC structure.

**Exposure estimation.** We estimated the proportion of workers exposed to wildfire PM_2.5_ for every day from 2010-2019, broken down by county and detailed SOC group. Our previous approach characterizing work-related heat exposure used Monte Carlo simulation for this estimation, with the probability of working outdoors specified as binomial distribution.^2^ However, our goal for this study was not to assess uncertainty in work-related wildfire smoke exposure, but rather the likely, central estimate of exposure. With enough Monte Carlo iterations, this simulated central estimate would be equivalent to multiplying the probability of working outdoors with the number of workers in a detailed SOC group. Using this modified approach and merging the ambient wildfire smoke levels on a given county-day, we estimated if outdoor workers are potentially exposed to smoke PM_2.5_ levels >9 µg/m^3^, the threshold defined by the US National Institute for Occupational Safety and Health (NIOSH) that requires employers to take control action to immediately protect workers from wildfire smoke.^1^

Thus, for each day, we estimated the proportion of workers exposed to wildfire PM_2.5_ >9 µg/m^3^ by detailed SOC group for each county in the contiguous US, accounting for differences in whether SOC groups work on weekends. We aggregated these detailed SOC estimates to estimate the proportion exposed by major SOC group for each county-day.

**Poststratification using major employment count data by county.** Lastly, we estimate the number of workers exposed to wildfire PM_2.5_ for each county through poststratification. The number of workers exposed is a function of the exposure probability (as estimated in the previous section) and the number of workers employed in each major occupational group. To achieve this, we extracted yearly breakdowns of the 22, non-military, 2010, major SOC employment counts by county from the 5-year, US American Community Survey (ACS) for poststratification of the major SOC-county estimates of exposure. This poststratification was done because we estimated the major SOC-county proportions based on metro/nonmetro-level, detailed SOC employment count data that will not be reflective of true county-level employment count estimations. The estimates were extracted using the `tidycensus` package in R. The estimates are further broken down by sex, so we aggregated the sex-specific estimates together. For males, we extracted variables 5-37 from Table C24010. For females, we extracted variables 41-73 from Table C24010.

We poststratified these daily-county-major SOC proportions using annual employment counts by major SOC group for each county. Estimates were not available before 2007, so we used the 2005-2009 ACS to represent the occupational composition of counties for 2006. Thus, for each day from 2006-2019, we estimated the number of outdoor workers potentially exposed to wildfire smoke PM_2.5_ for every county in the contiguous US. These estimates were aggregated both spatially (i.e., by state and nationally) as well as temporally (i.e., yearly and across 2006-2019).

**Limitations.** Several analytical decisions and assumptions were required to integrate multiple data sources, each with their own sources of bias and error. We only assessed workplace exposure among outdoor workers. Indoor workers can also be exposed to wildfire smoke, particularly if they work in an environment with poor ventilation. We chose not to consider indoor workplace exposures because there is substantial variability in poor indoor air quality with a lack of robust, publicly available information on workplace ventilation, making it difficult to incorporate this information into our estimates. While indoor workers are not the focus of our study, it is important to keep in mind that the total work-related population health burden of wildfire smoke is likely larger than our estimates that are limited to outdoor workers only. Another unmeasured factor that could underestimate the true population health burden is a lack of respiratory rate information. Workers who have to perform more physically demanding jobs will have a higher respiratory rate, increasing their exposure to wildfire smoke. However, this information is specific to an individual as it is driven by biological factors such as age and body weight, making it difficult to integrate into our estimates using publicly available data. Other information that could bias our estimates high or low in different contexts include the choice of wildfire smoke statistic in determining exposure (e.g., mean vs 95^th^ percentile), a lack of respirator use information (of which use is generally low), our determination of irregular working hours, and whether an individual works in the county they reside (e.g., long-haul truck drivers may be working in another state).

**Appendix B. Relationship between racial and ethnic composition and ambient and work-related exposure to wildfire PM_2.5_**

**Data sources.** All data used for modelling came from de-identified, publicly available, secondary data sources.

*Racial and ethnic composition.* For each year from 2006 to 2019, we extracted county-level estimates from 5-year, US American Community Survey data of the percentage of individuals who identify as (i) non-Hispanic White; (ii) non-Hispanic Black; and (iii) Hispanic. We excluded non-Hispanic Asian, American Indian/Alaskan Native, and Native Hawaiian/Pacific Islander individuals from this analysis since there are few few/no counties where a majority (>50%) of the county is comprised of these racial and ethnic subgroups. 5-year ACS data did not begin before the 2005-2009 interval, so we used 2005-2009 data to represent county-level racial and ethnic compositions in 2006.

*Ambient and work-related wildfire PM_2.5_ exposure.* We estimated the number of person-days exposed to work-related wildfire PM_2.5_ for a given county and year. For ambient exposure, we extracted daily, ambient wildfire PM_2.5_ estimates previously developed by Childs et al. between 2006-2019 for every county in the US.^3^ We also extracted yearly population counts from repeated, 5-year US American Community survey data to calculate county-level, population-weighted, yearly ambient wildfire PM_2.5_ levels. To mimic previous studies assessing ambient exposure inequities, which typically make use of all wildfire smoke days, we assumed that if any county-day had smoke PM_2.5_ >0 µg/m^3^, all residents in that county would be considered exposed. We summed up the number of person-days exposed across a given year, and this value reflected the ambient wildfire smoke exposure.

For work-related exposure, we summed up the number of person-days exposed to wildfire smoke across a year for a given county using the work-related wildfire smoke exposure metric constructed in this study (see **Appendix A** for details).

*Covariates.* To account for county differences in population sizes, we extracted annual county-level estimates from 5-year, US American Community Survey data of the total population from 2006 to 2019. Since we examined work-related exposure inequities, we controlled for the population unlikely to be employed using the number of residents <16 years old, residents >64 years old, and residents who are unemployed/in the armed forces/not in the labor force, all from yearly population counts by county from repeated, 5-year US American Community survey data between 2006-2019

**Model specifications.** We used Poisson, generalized additive models, as similarly used in a previous wildfire smoke-exposure inequity study^7^ and in our previous work-related heat inequity study,^2^ at the county-year level to assess which racial and ethnic groups may be disproportionately exposed to ambient or work-related wildfire smoke ($e$) for county $c$ and year $y$:

$$\log\left( E\left[ {exposure}_{ecy} \right] \right)= \beta_{0}+ns\left( \%nhWhite,df=4 \right)+ns\left( \%nhBlack,df=4 \right)+ns\left( \%Hispanic,df=4 \right)+{\beta_{1}\% Young}_{cy}+{\beta_{2}\% Old}_{cy}+{\beta_{3}\% Not\text{-}in\text{-}workforce}_{cy}+\beta_{y}+ns\left( {Longitude}_{c}+{Latitude}_{c},df=40 \right)+log({Population}_{cy})$$

where ${exposure}_{ecy}$ is the number of person-days exposed to ambient or work-related wildfire smoke; $\beta_{0}$ is the overall intercept; $ns\left( \%nhWhite,df=4 \right),$ $ns\left( \%nhBlack,df=4 \right)$, and $ns\left( \%Hispanic,df=4 \right)$ are the main effects of interest for the percent of individuals who are either non-Hispanic White, non-Hispanic Black, or Hispanic modelled using a natural spline with 4 degrees of freedom since non-linear relationships may over- or under-estimate racial and ethnic exposure inequities;^8^ ${\% Young}_{cy}$, ${\% Old}_{cy}$, ${\% Not\text{-}in\text{-}workforce}_{cy}$ are the percentage of individuals <16 years old, >64 years old, and those who are unemployed/in the armed forces/not in the labor force, respectively, to control for the population that is unlikely to be employed, and thus, exposed to wildfire smoke at work; $\beta_{y}$ is a categorical variable for year (2006-2019) to account for temporal autocorrelation; $ns\left( {Longitude}_{c}+{Latitude}_{c},df=41 \right)$ is each population-weighted, county centroid’s latitude/longitude (tensor product with 40 degrees of freedom) to account for spatial autocorrelation; ${Population}_{cy}$ is the total population of all ages, included as a population offset.

**Appendix C. Relationship between ambient wildfire PM_2.5_, workplace exposure, and all-cause mortality**

**Overview.** We empirically estimate the effect of wildfire smoke exposure on all-age, all-cause mortality within different strata of workplace smoke exposure (no or any workplace exposure) using a quasi-Poisson panel fixed effects model. Our goal was to replicate and expand upon a previous analysis of the wildfire smoke-mortality relationship;^9^ thus our analytical design and model specifications are informed by this previous study.

**Data sources.** All data used for modelling came from de-identified, publicly available, secondary data sources.

*Wildfire exposure data.* We extracted daily, population-weighted, ambient wildfire PM_2.5_ estimates previously developed by Childs et al. between 2006-2019 for every county in the contiguous US.^3^ Using the work-related wildfire smoke exposure rate constructed in this study (**Supplemental Figure S7**; see **Appendix A** for details), we classified county-years into two strata: (1) county-years with a rate of 0 were considered to have “no” workplace exposure and (2) those with a rate >0 had “any” workplace exposure.

Together, the ambient and work-related wildfire variables were combined into single, categorical variable to assess the joint effect. We constructed a ten-level categorical variable for whether annual-average, ambient wildfire smoke PM_2.5_ falls into the range of five bins (0-0.1, >0.1-0.25, >0.25-0.5, >0.5-0.75, >0.75 µg/m^3^) and the workplace exposure rate of wildfire smoke PM_2.5_ into two bins (“no”, “any”). While Qiu et al. (2025) assessed higher ambient exposures (e.g., >4 µg/m^3^), we were unable to due to a small sample size when stratifying by the work-related bins.

*Mortality outcome data.* We used publicly available mortality counts for 3,108 contiguous US counties between 2006-2019 from the National Center for Health Statistics.^10^ Counts were broken down by county and year, and included deaths from all-causes for all ages. Despite mortality counts being suppressed for county-years with <10 all-cause deaths, we used the publicly available mortality data rather than the restricted, full data, since Qiu et al. found a similar wildfire smoke-mortality relationship with either data source.^9^

*Covariates.* We extracted daily, mean air temperature data (in degrees Celsius) for each county in the US between 2006-2019 using previously published estimates from Spangler et al.^11^ We calculated average wet-bulb globe temperature for each county-year. We also extracted yearly, mean precipitation data (in inches) for each county in the US between 2006-2019 made publicly available by the US National Oceanic and Atmospheric Administration (NOAA) National Centers for Environmental Information.^12^

**Model specifications.** We use a similar quasi-Poisson, panel fixed effects regression model as previously used in a wildfire smoke-mortality study,^9^ at the county-year level. We assess the joint effect of average, ambient wildfire PM_2.5_ levels and workplace exposure on the percent change all-cause mortality rates for county $c$, state $s$, and year $y$:

$$\log\left( E\left[ {deaths}_{csy} \right] \right)= \beta_{c}+\beta_{sy}+\sum_{w=1}^{2} \sum_{a=1}^{5} \left( \beta_{wa} {Work\text{-}Ambient Exposure Bins}_{csy}^{wa} \right)+ns\left( {Temperature}_{csy}, df=5 \right)++ns\left( {Precipitation}_{csy}, df=5 \right)+log({Population}_{csy})$$

where ${deaths}_{csy}$ is the number of all-cause deaths; $\beta_{c}$ is the county-specific intercept that accounts for any county-specific, time-invariant factors that could be associated with both smoke exposure and mortality (e.g., income); $\beta_{sy}$ is the state-year specific intercept to control for spatial confounders that vary across state and across time (e.g., policy); $\beta_{wa}$ are the main coefficients of interest for categories of the joint exposure to ambient and workplace wildfire smoke; ${Work\text{-}Ambient Exposure Bins}_{csy}^{wa}$ is a ten-level categorical variable for whether annual-average, ambient wildfire smoke PM_2.5_ falls into the range of bin $a$ (0-0.1, >0.1-0.25, >0.25-0.5, >0.5-0.75, 0.75+ µg/m^3^) and the workplace exposure rate of wildfire smoke PM_2.5_ is 0 or >0 exposures per 10,000 workers, where county-years with both the lowest ambient levels are the reference category for each workplace exposure rate stratum; $ns\left( {Temperature}_{csy}, df=5 \right)$ and $ns\left( {Precipitation}_{csy}, df=5 \right)$ are natural spline terms with 5 degrees of freedom to account for the time-varying effect of wet-bulb globe temperature and precipitation on mortality; and ${Population}_{csy}$ is the total population of all ages, included as a population offset. The quasi-Poisson model accounts for overdispersion of mortality counts. The 95% confidence intervals of the coefficients were estimated with 500 bootstrap iterations of the model, taking the 2.5^th^ and 97.5^th^ percentile. Modelling was performed using the `feglm` function in the `fixest` R package.

Our modelling approach here examines the effect of wildfire smoke exposure bins on all-cause mortality using within-county variation over time, which removes confounding bias due to time-invariant factors that are unlikely to vary across counties in this time period (e.g., rurality, educational attainment, general socioeconomic conditions). Additionally, our approach accounts for factors that differ within a state in a given year using state-year fixed effects (e.g., wildfire smoke regulation) and for potential time-varying confounding bias between wildfire smoke and mortality due to temperature or precipitation. While others have reported effect estimates at higher, annual average, ambient wildfire smoke concentrations (e.g., >5 µg/m^3^), we truncated our estimates to >0.75 µg/m^3^ to ensure sufficient sample size when stratifying between county-years by workplace exposure to wildfire smoke. To understand the effects of this truncation, we replicate previous estimates^9^ using the full range of ambient concentrations and compare to estimates with truncated concentrations (see **Replication and** **Sensitivity Analysis** below).

We additionally explored how the model coefficients might differ if information on workplace exposure to wildfire smoke is not incorporated. We ran a similar quasi-Poisson regression model with the same model specifications and covariates as the model above, but where our main exposure of interest is instead the five, annual-average, ambient wildfire smoke PM_2.5_ bins only. Specifically, we modeled:

$$\log\left( E\left[ {deaths}_{csy} \right] \right)= \beta_{c}+\beta_{sy}+\sum_{a=1}^{6} \left( \beta_{a} {Ambient Exposure Bins}_{csy}^{a} \right)+ns\left( {Temperature}_{csy}, df=5 \right)+ns\left( {Precipitation}_{csy}, df=5 \right)+log({Population}_{csy})$$

where instead the joint effect of ambient and wildfire smoke exposure ($\beta_{wa}$) are the main coefficients of interest, only ambient wildfire smoke is ($\beta_{a}$).

**Replication and sensitivity analysis.** We compared our model results to those previously reported by Qiu et al. to investigate the robustness of our findings, who similarly ran a quasi-Poisson, fixed effects regression model between ambient wildfire PM2.5 concentrations and all-cause mortality at the county-year level from 2006-2019.^9^ We first re-ran the same model specified by Qiu et al. to examine whether our model estimates are similar and reliable, given we used slightly different data sources:

$$\log\left( E\left[ {deaths}_{csy} \right] \right)= \beta_{c}+\beta_{sy}+\sum_{a=1}^{9} \left( \beta_{a} {Ambient Exposure Bins}_{csy}^{a} \right)+ns\left( {Temperature}_{csy}, df=5 \right)+ns\left( {Precipitation}_{csy}, df=5 \right)+log({Population}_{csy})$$

where the only difference between Qiu et al.’s ambient wildfire smoke model and our is the number of ambient exposure bins (Qiu et al. further binned intervals >1: 1-2, 2-3, 3-4, 4+ µg/m^3^). We compared the effect estimates originally published by Qiu et al. to (1) our model using Qiu et al.’s model specifications and (2) our model with ambient exposure bins truncated at >1 µg/m^3^ (**Supplemental Figure S8**). We did not observe substantial differences and the exposure-response function maintained a similar shape, although the effect estimates from our data were slightly attenuated towards the null.

Around 1% of county-years (451 of 43,469 county-years) in this analysis had suppressed all-cause mortality counts due to low counts (<10 deaths). The model presented in the main text imputed these suppressed values with the midpoint (4.5 deaths). To examine how the effect estimates could be influenced by the choice in dealing with the suppressed values, we compared the main model to three models where the county-years with suppressed values were either imputed with 0 deaths, 9 deaths, or were dropped from the analysis, or where an entire county with any county-years suppressed was dropped. In general, the method of handling the suppressed values did not alter the effect estimates (**Supplemental Figure S9**).

**Supplementary Table S1.** NIOSH exposure control categories (ECC) by Air Quality Index groupings as defined in Table 4-1 in the draft 2024 NIOSH Hazard Review “Wildland Fire Smoke Exposure Among Farmworkers and Other Outdoor Workers”.^1^

| **NIOSH ECC** | **PM_2.5_**  (µg/m^3^) | **Exposure**  **definitions** |
| --- | --- | --- |
| 1 | 0.0 to 9.0 | Unexposed |
| 2 | 9.1 to 35.4 | Exposed |
| 3 | 35.5 to 55.4 | Exposed |
| 4 | 55.4 to 125.4 | Exposed |
| 5 | 125.5 to 225.4 | Exposed |
| 6 | 225.5+ | Exposed |

**Supplemental Table S2.** Percent change (95% confidence interval) between average, ambient smoke PM2.5 and all-cause mortality rates between 2006-2019 among all counties (pooled), among counties with no (0 exposure-days per 10,000 workers), or among counties with any (>0 exposure-days per 10,000 workers) workplace exposure from panel fixed effects quasi-Poisson regression models of county-years. Effects visualized in Figure 2b.

| **Wildfire smoke PM_2.5_** | **Pooled** | | **No work exposure** | | **Work exposure** | |
| --- | --- | --- | --- | --- | --- | --- |
|  | **N** | **Effect** | **N** | **Effect** | **N** | **Effect** |
| <0.1 µg/m³ | 4,604 | 0% (ref) | 4,092 | 0% (ref) | 512 | 0% (ref) |
| 0.1-0.25 µg/m³ | 12,313 | 0.4% (0.0, 0.7) | 7,442 | 0.2% (-0.1, 0.6) | 4,871 | 1.2% (0.5, 2.0) |
| 0.25-0.5 µg/m³ | 14,552 | 0.6% (0.1, 1.0) | 4,012 | 0.7% (0.2, 1.2) | 10,540 | 1.5% (0.6, 2.2) |
| 0.5-0.75 µg/m³ | 6,798 | 0.5% (0.0, 1.1) | 518 | -0.2% (-1.1, 0.5) | 6,280 | 1.5% (0.6, 2.4) |
| 0.75+ µg/m³ | 5,202 | 0.7% (0.0, 1.5) | 199 | -0.5% (-1.7, 0.9) | 5,003 | 1.7% (0.7, 2.7) |

**Supplemental Table S3.** List of each variable used to construct work-related wildfire smoke exposure rates across three steps (Step 1: Data integration; Step 2: Exposure estimation; and Step 3: Poststratification).

| **Step** | **Variable** | **Data source** | **Temporal scale** | **Spatial scale** | **Data description** | **Variable definition** |
| --- | --- | --- | --- | --- | --- | --- |
| 1 | Wildfire smoke levels | Childs et al. (2022)^3^ | Daily, 2000-2020 | County | Ambient wildfire PM_2.5_ levels (estimated in µg/m^3^) for every county in the contiguous United States each day from January 1, 2000 to December 31, 2020 | Classified into six categories (0.0-9.0; 9.1-35.4; 35.5-55.4; 55.4-125.4; 125.5-225.4; 225.5+ µg/m^3^) |
| 1, 2 | Outdoor work | O*NET v24.1^4^ | Decadal, 2010-2019 | Nationwide | An estimate of the “Frequency Required to Work Outdoors, Exposed to Weather” work context variable (element ID #4.C.2.a.1.c), reflecting “How often does this job require working outdoors, exposed to all weather conditions?” (scale of 0-100) for every detailed SOC group | Converted to probability of working outdoors on a given day (0-100%) |
| 1 | Irregular work hours | O*NET v24.1^4^ | Decadal, 2010-2019 | Nationwide | An estimate of the “Duration of Typical Work Week” work context variable (element ID #4.C.3.d.8), reflecting “Number of hours typically worked in one week” (scale of 0-100) for every detailed SOC group | If either the first value was >50 or the second value was >25, detailed SOC group was considered as working irregular hours |
|  |  | O*NET v24.1^4^ | Decadal, 2010-2019 | Nationwide | An estimate of the “Work Schedules” work context variable (element ID #4.C.3.d.4), reflecting “How regular are the work schedules for this job?” (scale of 0-100) for every detailed SOC group |  |
| 1 | Detailed employment counts | 2019 BLS OEWS^6^ | Decadal, 2010-2019 | Metropolitan/  Nonmetropolitan area | Metropolitan/nonmetropolitan area employment counts of 840 detailed SOC groups from 2010-2019 | Number of civilian, non-military, employed individuals |
| 3 | Major employment counts | US American Community Survey | Yearly, 2010-2019 | County | County employment counts of 22 major SOC groups from 2010-2019, aggregated from sex-specific estimates. For males, we extracted variables 5-37 from Table C24010. For females, we extracted variables 41-73 from Table C24010 | Number of civilian, non-military, employed individuals |

**Supplemental Figure S1.** Annual average ambient wildfire smoke PM_2.5_ levels (µg/m^3^) among county-years with no estimated work-related exposure to wildfire smoke. States with white border have occupational regulations on wildfire smoke as of 2025.

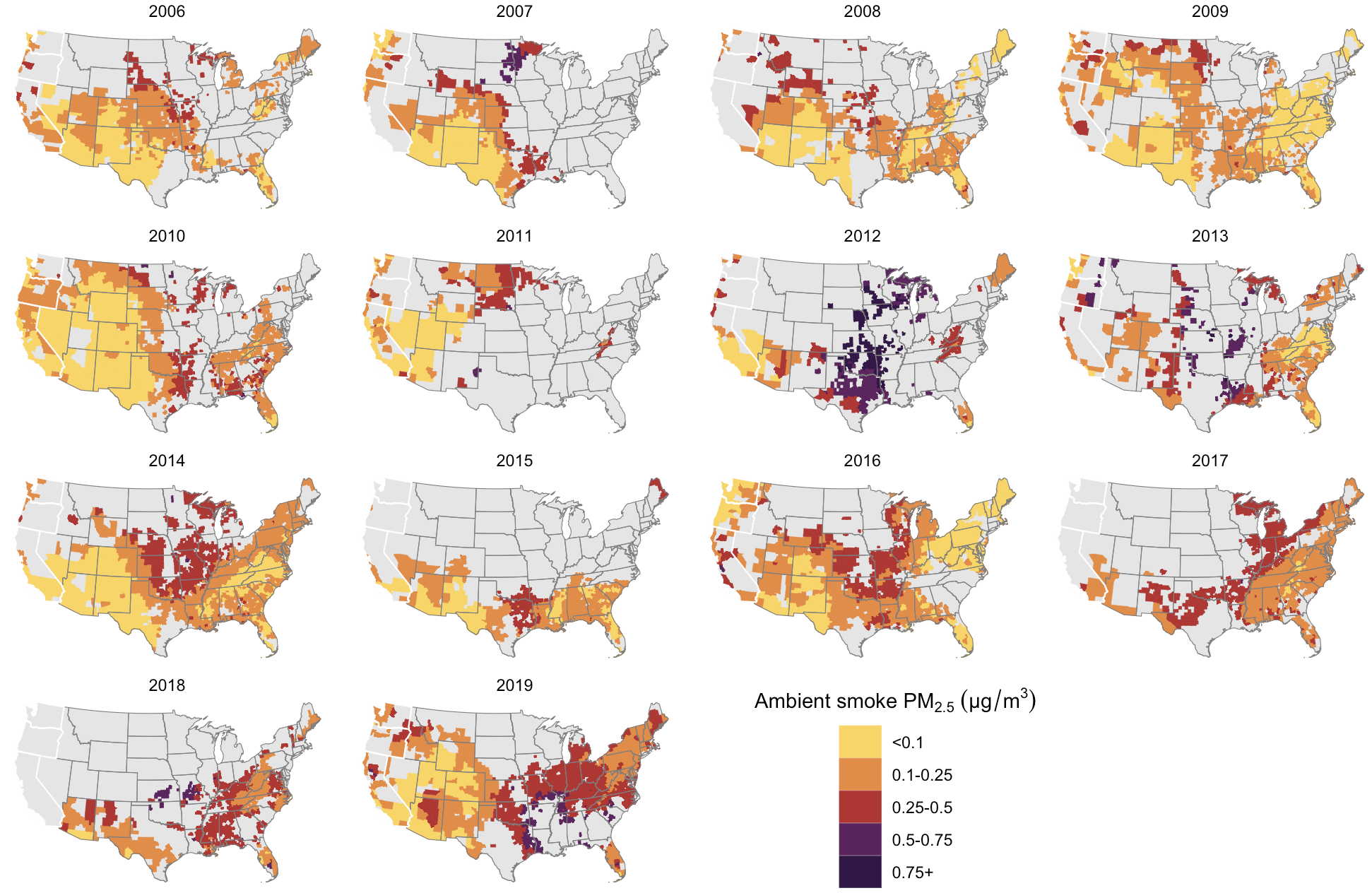

**Supplemental Figure S2.** Annual average ambient wildfire smoke PM_2.5_ levels (µg/m^3^) among county-years with any work-related exposure to wildfire smoke. States with white border have occupational regulations on wildfire smoke as of 2025.

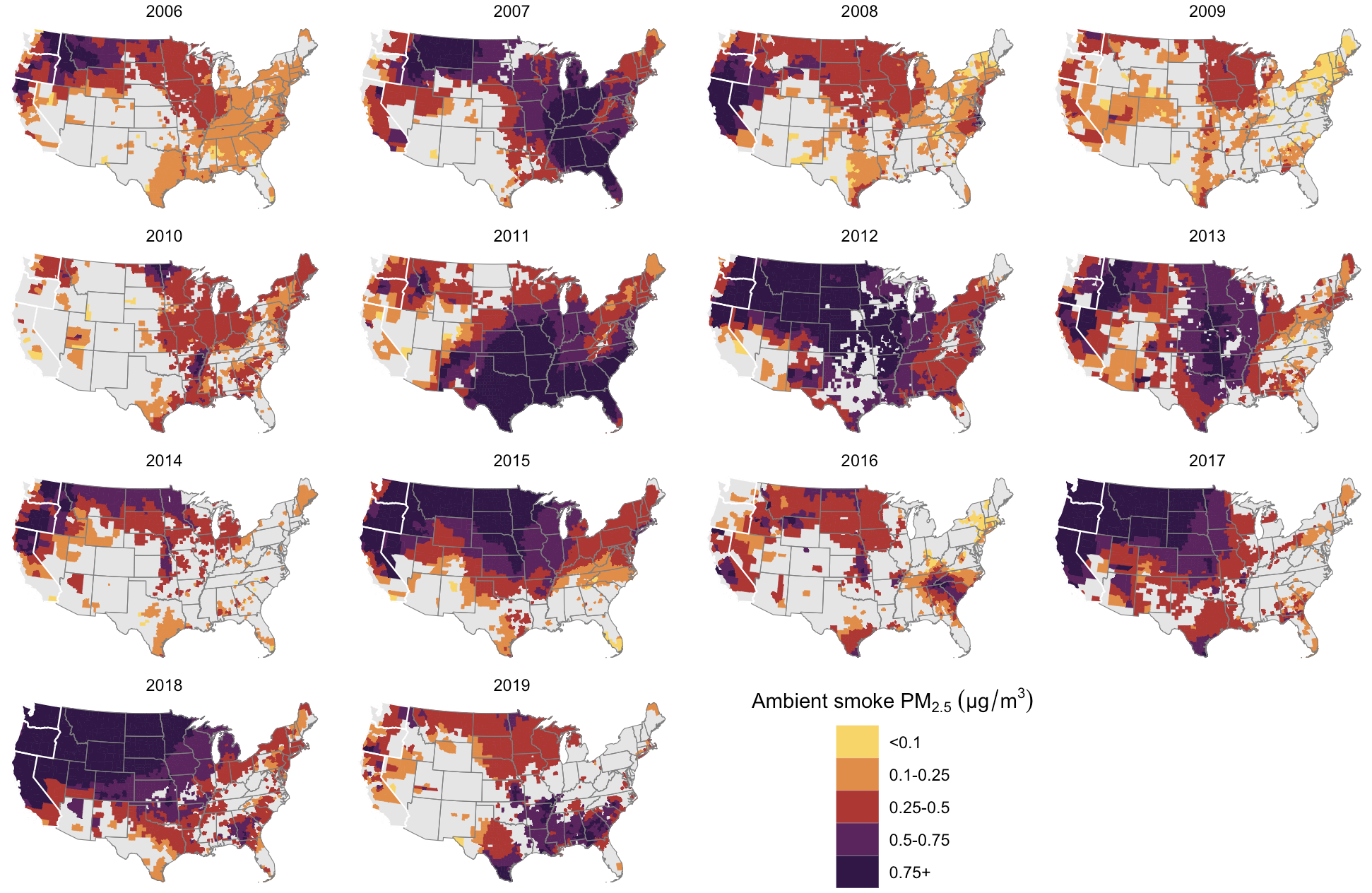

**Supplemental Figure S3.** Flowchart illustrating data inputs, statistical methodology, and output of the workplace exposure rate per 1,000 workers. BLS OEWS: Bureau of Labor Statistics Occupational Employment and Wage Statistics; O*NET: Occupational Information Network; US ACS: United States American Community Survey.

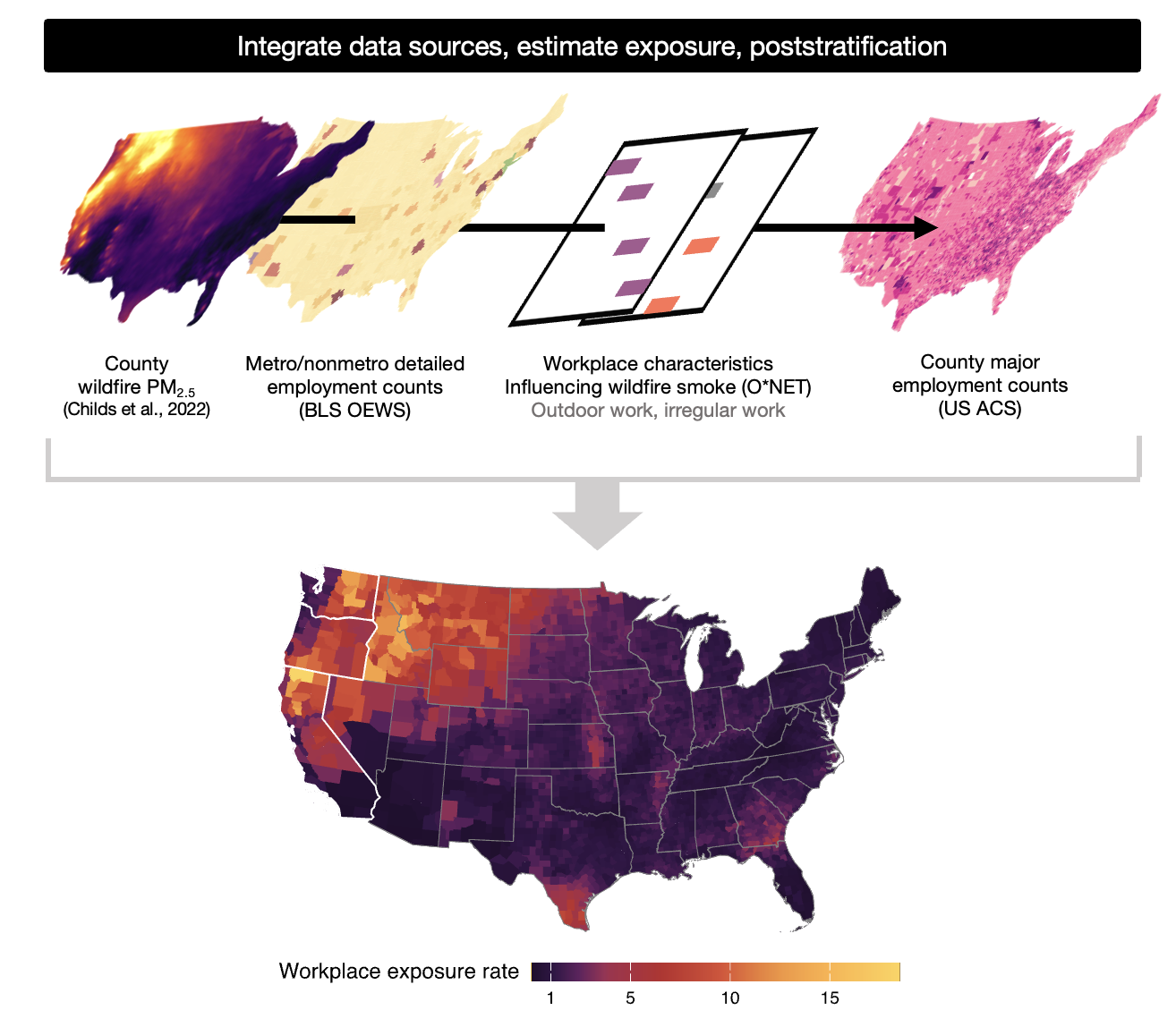

**Supplemental Figure S4.** Histogram by detailed SOC group (n=760) of the proportion of work-year outdoors

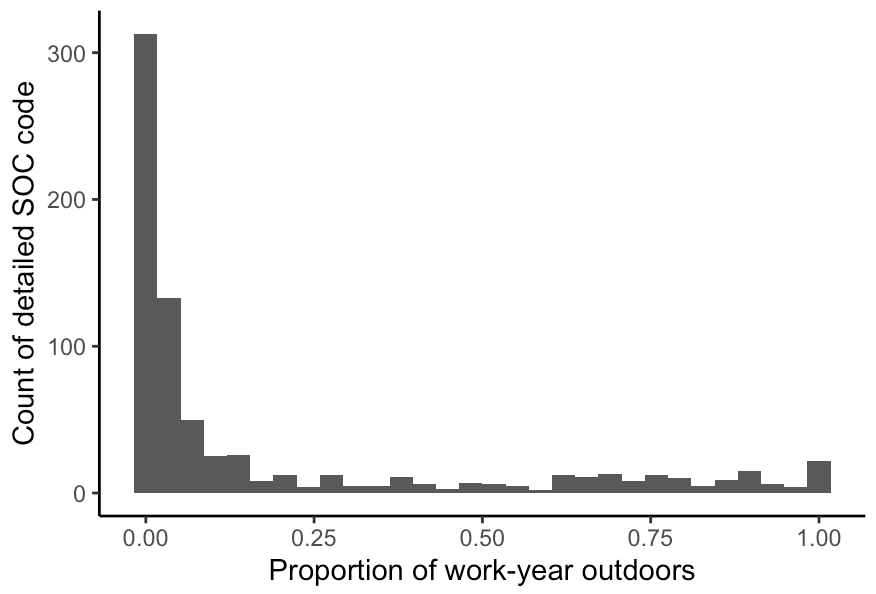

**Supplemental Figure S5.** Yearly, likely number of outdoor workers by county. States with white border have occupational regulations on wildfire smoke as of 2025.

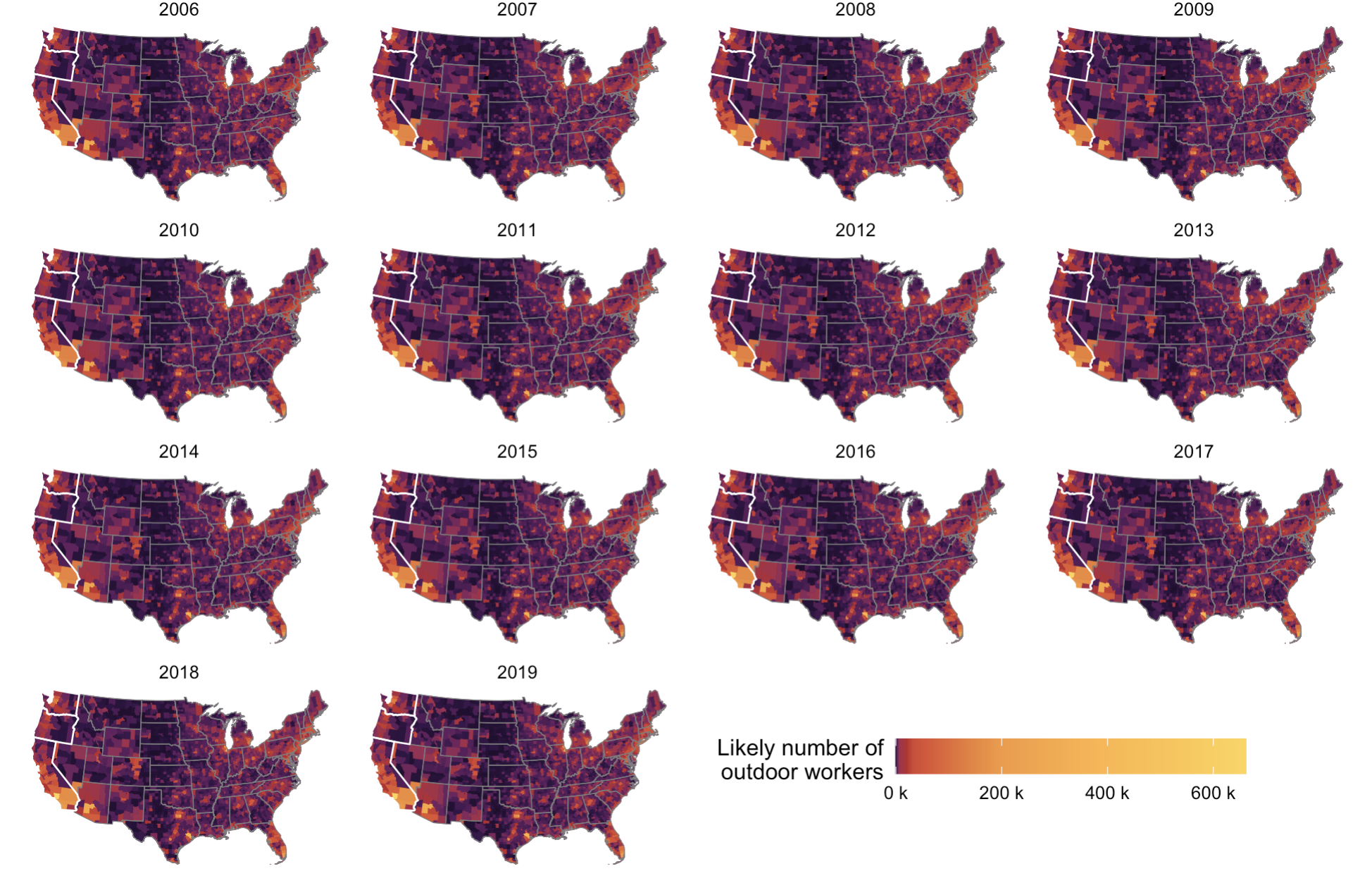

**Supplemental Figure S6.** Bar chart by detailed SOC group (n=760) of irregular working hours and could work on the weekends (value = 1)

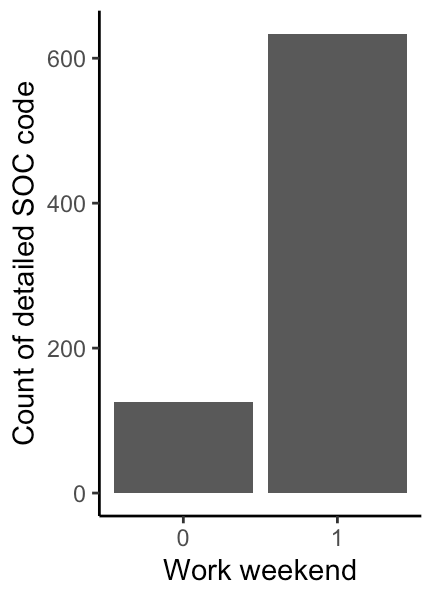

**Supplemental Figure S7.** Yearly, average rate of work-related wildfire PM_2.5_ exposure ≥9 µg/m^3^ per 10,000 workers by county. States with white border have occupational regulations on wildfire smoke as of 2025.

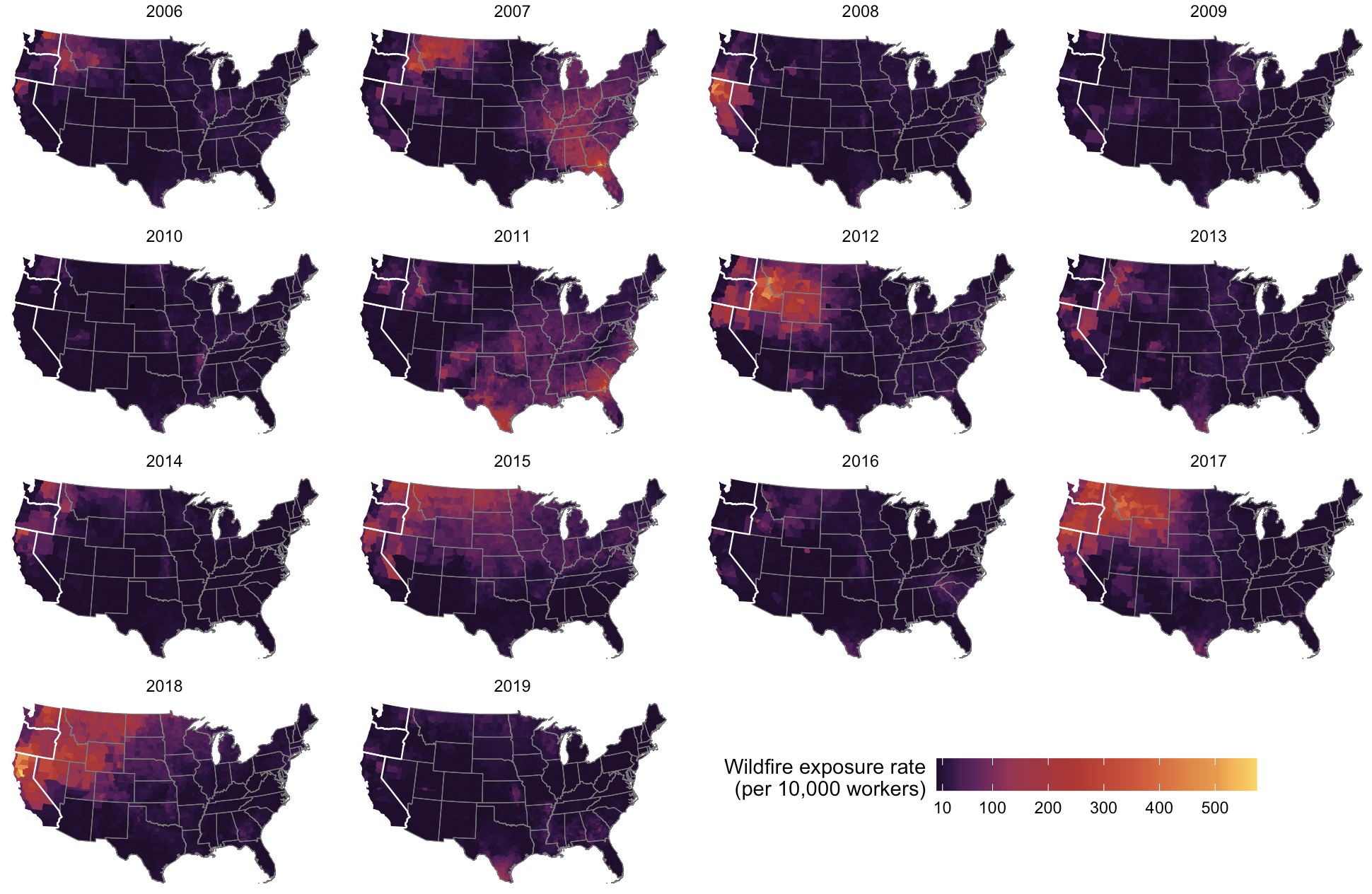

**Supplemental Figure S8.** Sensitivity analysis comparing original effect estimates (top to bottom) reported by Qiu et al. (using restricted-use mortality data without suppressed mortality counts), replicating Qiu et al.’s model (using publicly available mortality data with suppressed mortality counts and different data sources for model covariates), and truncating the replicated model to >0.75 µg/m^3^.

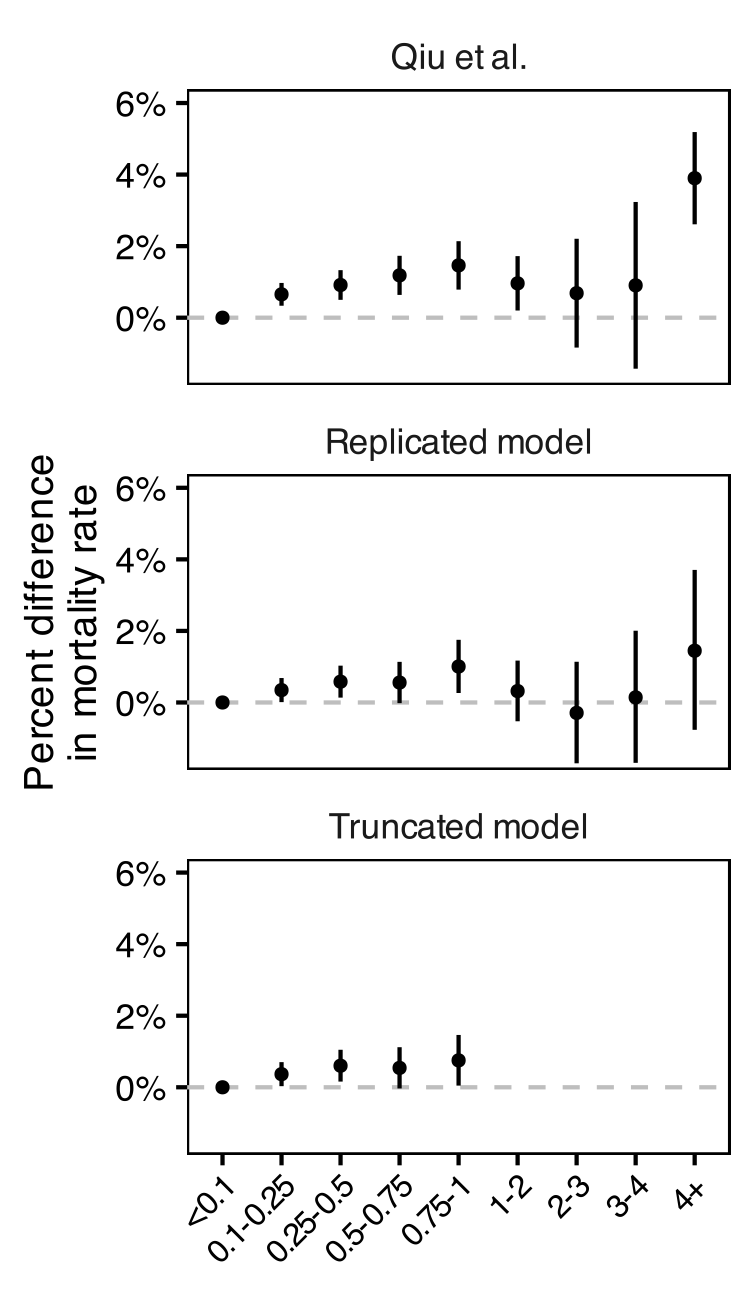

**Supplemental Figure S9.** Sensitivity analysis comparing effect estimates from differences in handling suppressed mortality values among county-years with <10 deaths. Top to bottom: imputing suppressed values to 4.5 (mid point of 0-9 deaths; main model); imputing suppressed values to 0; imputing suppressed values to 9; dropping county-years with suppressed values.

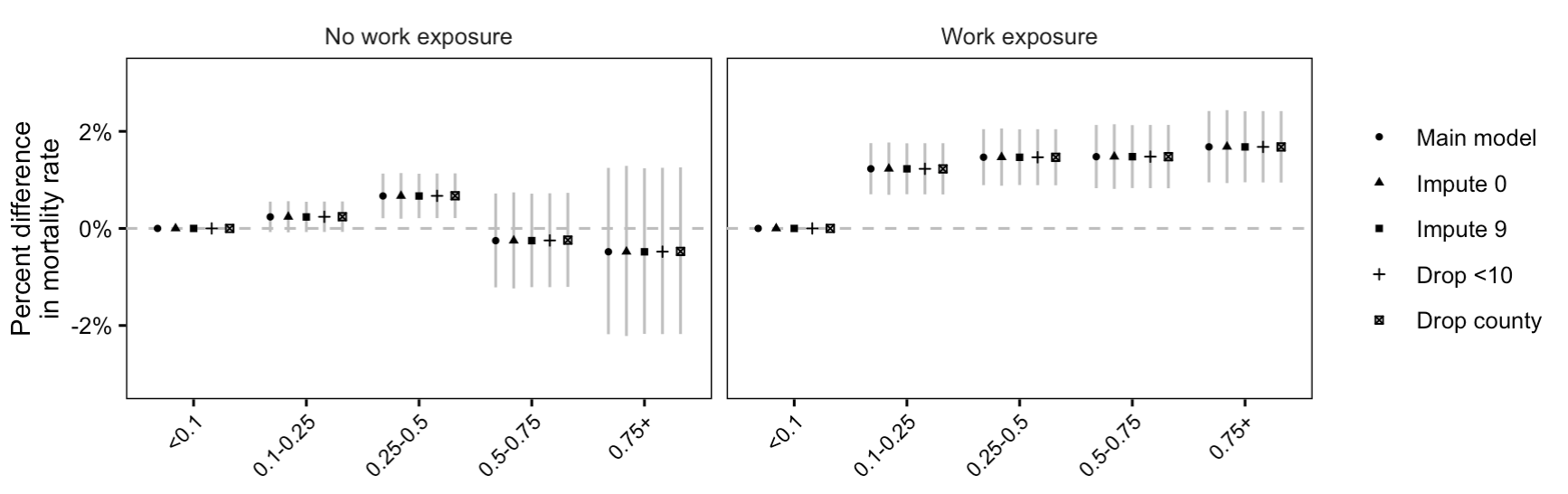
